# Estimating Variation of Covid-19 ‘infection’ in the Population: Results from Understanding Society’s (UKHLS) first monthly covid-19 survey

**DOI:** 10.1101/2020.07.22.20159806

**Authors:** Richard Breen, John Ermisch

## Abstract

The analysis in this paper uses the new *Understanding Society COVID-19* survey. The key advantage of these data is that they allow us to examine infection rates for people with particular characteristics. We study how reported symptoms vary in the population and relate reported symptoms to a positive Covid-19 test in the small sample in the survey who were tested. Combining these probabilities we find that the chances of infection increase with a person’s education level, are lower and declining with age among those aged over 55, and were higher in the West Midlands and London and lower in the North East than in the rest of the country, and tended to increase with regional population density. There is also evidence that the infection rate was lower among those of a Caribbean origin. A suitably cautious estimate of the mean infection rate is that, during the period up to the end of April 2020, it was between 2% and 8%, with a central rate of about 5%.

---

The aim of the paper is to examine how the probability of Covid-19 infection varies within the UK population. It uses the new *Understanding Society COVID-19* survey to do so. In the main analysis we find that the mean cumulative infection rate in the UK population living in private households was 0.047 during the period up to the end of April 2020, with a 95% confidence interval of between 0.024 and 0.070. The key advantage of these data is the ability to examine infection rates for people with particular characteristics. The analysis indicates that the infection rate based on reported symptoms (and the relationship between reported symptoms and a positive test) increases with a person’s education level, is lower and declining with age among those aged over 55, was higher in the West Midlands and London and lower in the North East than in the rest of the country, and tended to increase with regional population density. There is also evidence that the infection rate was lower among those of a Caribbean origin.

## The data

The COVID-19 survey is a monthly web-based survey on the experiences and reactions of the UK population to the COVID-19 pandemic.^1^ The 20 minute questionnaire includes core content repeated monthly to track changes through the pandemic, as well as rotating content. The ‘active’ sample includes everyone in households who have participated in at least one of the last two waves of data collection in Understanding Society. Among these, all household members who were aged 16 or over in April 2020 were invited to join the COVID-19 study. 42,330 sample members were sent a letter inviting them to participate in the study and fieldwork was carried out during 24-30 April. 16,379 respondents fully completed the survey and 17,452 responded partially (i.e. they at least completed the core coronavirus illness module), yielding full and partial response rates of 38.7 and 41.2%, respectively.

Because of the low level of testing during March and April 2020, measures of Covid-19 infection from these data must be mainly based on reported symptoms, not positive tests, although the paper uses the survey’s available test data to convert symptoms into infection probabilities. The primary measures used are the response to the question: *Have you experienced symptoms that could be caused by coronavirus (COVID-19)?* The response is recoded ‘no’ if they experienced none of the 10 symptoms listed on the questionnaire, and we narrow the focus further onto the following four common Covid-19 symptoms: 1. High temperature; 2. A new continuous cough; 3. Loss of sense of smell or taste.; and 4. Fatigue. The first three are the most common symptoms according to Public Health England (PHE), and the last was also common in a recent study by Sudre et al. (*Symptom clusters in Covid19: A potential clinical prediction tool from the COVID Symptom study app*, June 2020).^21^

We use a probabilistic measure of infection based on the number of these four symptoms that a person reports. The distribution is shown in Table 1.

**Table 1:** Percentage of respondents with exact number of 4 common Covid-symptoms*

|  | No symptom | 1 symptom | 2 symptoms | 3 symptoms | 4 symptoms |
| --- | --- | --- | --- | --- | --- |
| Per cent (weighted) | 89.1 | 3.9 | 3.6 | 2.7 | 0.8 |
| Unweighted N | 15,445 | 706 | 662 | 478 | 153 |

\*High temperature; a new continuous cough; loss of sense of smell or taste; fatigue.

Table 2 shows the incidence of the 4 symptoms by the number of symptoms reported. In each sum category, fatigue is the most likely to be reported. Loss of taste or smell is least likely to be among the four symptoms.

**Table 2:** Incidence (%) of 4 common Covid-symptoms by number of symptoms

| Symptoms | 1 symptom | 2 symptoms | 3 symptoms | 4 symptoms |
| --- | --- | --- | --- | --- |
| High temperature | 14.5 | 47.1 | 82.4 | 100 |
| New continuous cough | 33.6 | 52.9 | 78.5 | 100 |
| Loss of taste or smell | 9.4 | 19.9 | 45.2 | 100 |
| Fatigue | 42.6 | 80.1 | 93.9 | 100 |

We have sparse evidence from the survey to validate the degree to which the chances of being tested positive for the coronavirus increase with the number of the four symptoms. Of those reporting any of the four, only 6% were tested for coronavirus, with the percentage tested increasing with the number of symptoms reported (bottom row of Table 3). Amongst those tested, 19% of tests were either inconclusive or the people were still waiting for the results. With these limited data (N=147), we find that the percentage testing positive increases with the number of symptoms, as Table 3 indicates. In a simple linear regression, each additional symptom raises the probability of a positive test by 0.164 (SE=0.022).^3^

**Table 3:**
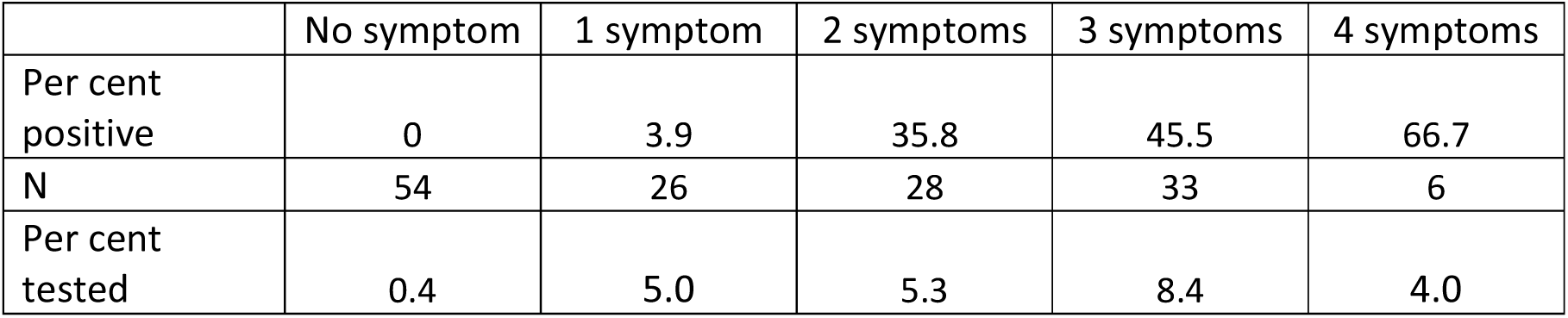
Percentage of tested respondents with positive result by number of 4 common Covid-symptoms (must have had received test results and had a conclusive test)

## Methods

In the probabilistic approach used here we assume that there is a latent index for ‘infection’ which is increasing in the number of the four symptoms reported, and which is a function of various attributes of the respondent (e.g. age). For any person with a given set of attributes *X* we can estimate the probability of having *n* of these four symptoms, *P*(*n*|*X*). Then if the probability of testing positive given *n* is *q*(*n*), the probability of infection is P(*infection*|*X)*= *q*(*n*)*P*(*n*|*X*). For example, if *q*(*n*) is given by a logit regression model linear in *n*, then, together with our model for the number of symptoms reported, we obtain an estimate of P(*infection*|*X)*.

## Results for P(n|X)

The symptoms data for each respondent in the covid-19 survey are matched to the last available wave of Understanding Society in which they participated. For 97% of the sample, this was wave 9, and for another 3 % it was wave 8. From these matched data, and from the Covid-19 survey, we can derive a large number of social, demographic, economic and health variables.

We assume that *P*(*n*|*X*) is given by an ordered model based on the standard normal distribution; i.e. an ordered probit model.^4^ The variables *X* affect the latent index, and from that association and the standard normal distribution function we can infer the association between *P*(*n*|*X*) and the following individual or household variables: age group, gender, ethnic group, region, household density (persons per room), urban (vs rural) area, highest education level, equivalent household income quintile based on multi-year averages of income,^5^ and whether or not the person lived in a household which owned their house outright. The analysis also controls for a number of reported health conditions which may also produce Covid-19 symptoms, including asthma, angina, emphysema, chronic bronchitis, COPD, high blood pressure and coronary heart disease. Other than asthma and chronic bronchitis, the coefficients of these health conditions are small.

The coefficients in Table 4 indicate the impact of the variable (relative to its base category) on the underlying latent variable, which has implications for the probabilities of being in each of the four categories. Those that stand out as being at least twice their standard error are indicated in bold type.

**Table 4:**
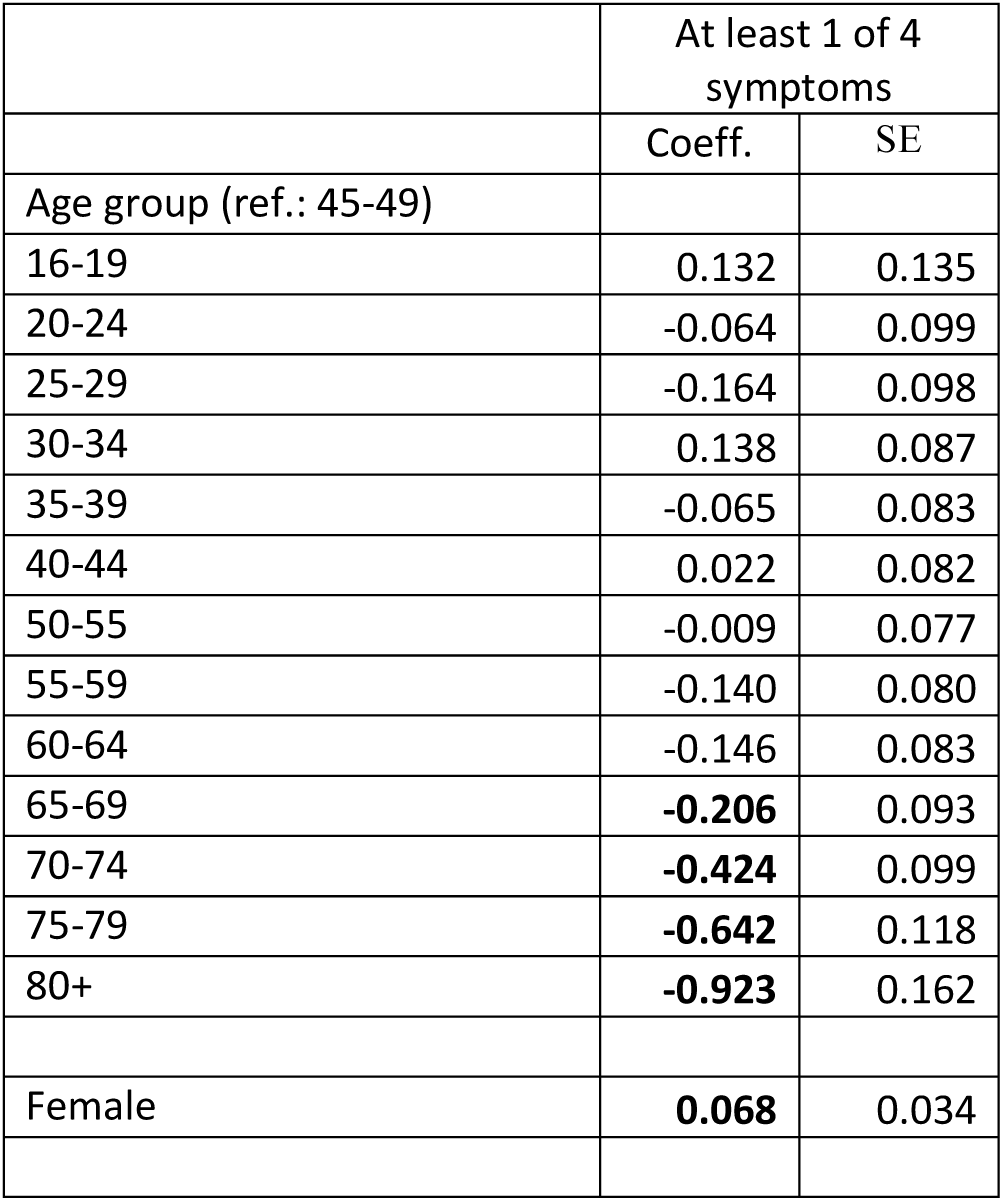
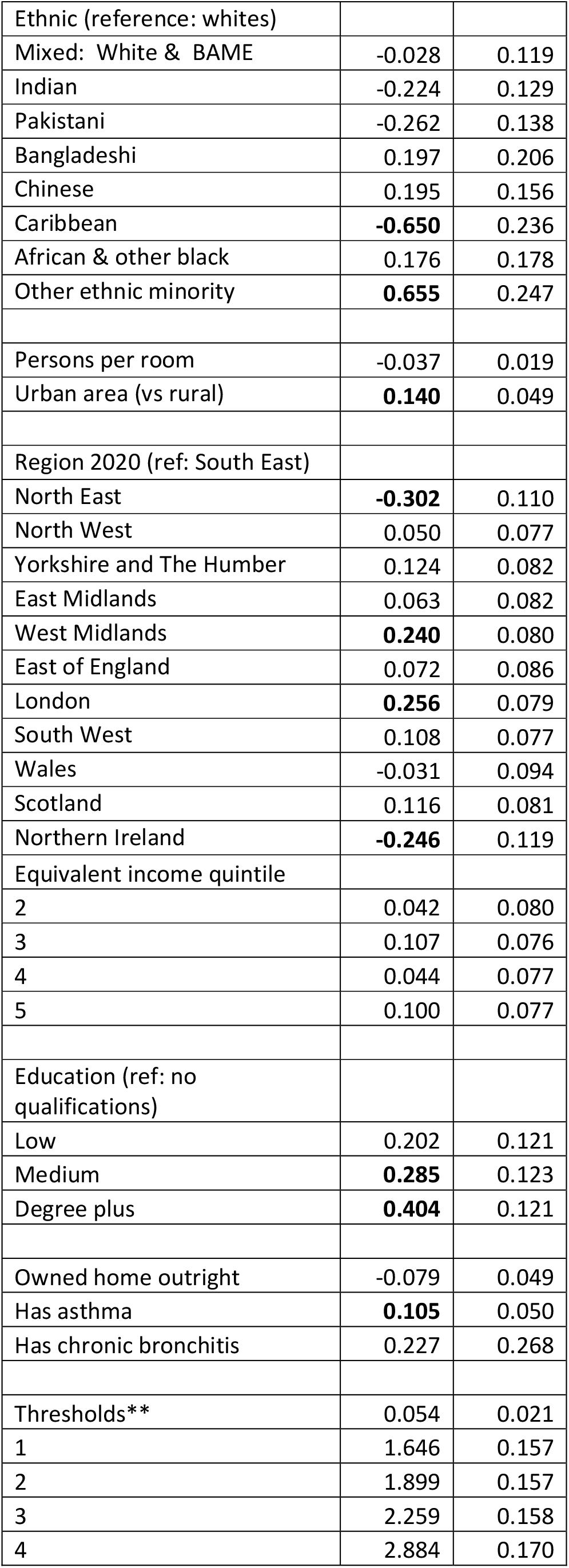
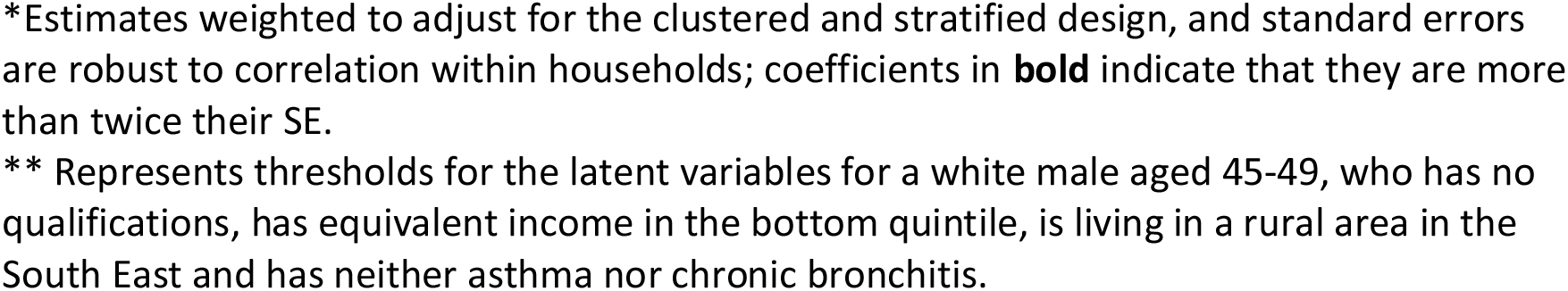
Ordered probit coefficient estimates for the sum of 4 symptoms*

|  | At least 1 of 4 symptoms |  |
| --- | --- | --- |
|  | Coeff. | SE |
| Age group (ref.: 45-49) |  |  |
| 16-19 | 0.132 | 0.135 |
| 20-24 | -0.064 | 0.099 |
| 25-29 | -0.164 | 0.098 |
| 30-34 | 0.138 | 0.087 |
| 35-39 | -0.065 | 0.083 |
| 40-44 | 0.022 | 0.082 |
| 50-55 | -0.009 | 0.077 |
| 55-59 | -0.140 | 0.080 |
| 60-64 | -0.146 | 0.083 |
| 65-69 | **-0.206** | 0.093 |
| 70-74 | **-0.424** | 0.099 |
| 75-79 | **-0.642** | 0.118 |
| 80+ | **-0.923** | 0.162 |
| Female | **0.068** | 0.034 |
| Ethnic (reference: whites) |  |  |
| Mixed: White & BAME | -0.028 | 0.119 |
| Indian | -0.224 | 0.129 |
| Pakistani | -0.262 | 0.138 |
| Bangladeshi | 0.197 | 0.206 |
| Chinese | 0.195 | 0.156 |
| Caribbean | **-0.650** | 0.236 |
| African & other black | 0.176 | 0.178 |
| Other ethnic minority | **0.655** | 0.247 |
| Persons per room | -0.037 | 0.019 |
| Urban area (vs rural) | **0.140** | 0.049 |
| Region 2020 (ref: South East) |  |  |
| North East | **-0.302** | 0.110 |
| North West | 0.050 | 0.077 |
| Yorkshire and The Humber | 0.124 | 0.082 |
| East Midlands | 0.063 | 0.082 |
| West Midlands | **0.240** | 0.080 |
| East of England | 0.072 | 0.086 |
| London | **0.256** | 0.079 |
| South West | 0.108 | 0.077 |
| Wales | -0.031 | 0.094 |
| Scotland | 0.116 | 0.081 |
| Northern Ireland | **-0.246** | 0.119 |
| Equivalent income quintile |  |  |
| 2 | 0.042 | 0.080 |
| 3 | 0.107 | 0.076 |
| 4 | 0.044 | 0.077 |
| 5 | 0.100 | 0.077 |
| Education (ref: no qualifications) |  |  |
| Low | 0.202 | 0.121 |
| Medium | **0.285** | 0.123 |
| Degree plus | **0.404** | 0.121 |
| Owned home outright | -0.079 | 0.049 |
| Has asthma | **0.105** | 0.050 |
| Has chronic bronchitis | 0.227 | 0.268 |
| Thresholds** | 0.054 | 0.021 |
| 1 | 1.646 | 0.157 |
| 2 | 1.899 | 0.157 |
| 3 | 2.259 | 0.158 |
| 4 | 2.884 | 0.170 |

\*Estimates weighted to adjust for the clustered and stratified design, and standard errors are robust to correlation within households; coefficients in **bold** indicate that they are more than twice their SE.
\*\* Represents thresholds for the latent variables for a white male aged 45-49, who has no qualifications, has equivalent income in the bottom quintile, is living in a rural area in the South East and has neither asthma nor chronic bronchitis.

### These patterns stand out

**Age and Sex**: persons age 55 and older have fewer symptoms than those aged 45-49 (the reference group), and their chances of being infected decline with age. This pattern probably reflects policies and guidance promoting self-sheltering among the elderly. In light of the high death rates from Covid-19 among people of these ages, this finding strongly suggests that they are less likely to be infected but more likely to die if infected. Women report more slightly more symptoms on average.

**Ethnic origin**: compared to whites, people of Caribbean origin are much *less* likely to have many symptoms. But, like older people, the higher death rates for people with a BAME background suggest that those of Caribbean origin are less likely to become infected but much more likely to die if infected.

**Location**: Living in an urban area is associated with a larger number of symptoms, and people from the West Midlands and London are likely to report more symptoms, while people from the North East are less likely to do so.

**Socio-economic status**: the number of reported symptoms increases with a person’s education level, and it is much higher for degree educated people. This association may reflect differences in recalling experience of symptoms as well as their incidence. All else equal, the number of symptoms is lower among people living in a house owned outright.

A cross-validation analysis was carried out. First, a sample of 11,000 observations was drawn for estimation of the parameters. We then compare the predictions of the distribution of the sum of the four symptoms from the model using the estimated parameters with the actual outcome for another sample of 4,738 people. The two distributions are compared in Table 5. We cannot reject the hypothesis that the distribution of actual outcomes in the sample is the same as the predicted distribution: the Kolmogorov– Smirnov test statistic is K-S=0.007 (0.05 critical value =0.020).

**Table 5:** Predicted and Actual Outcomes in Post-estimation Sample

| Symptoms | Predicted | Actual | Difference |
| --- | --- | --- | --- |
| 0 | 4,195 | 4,158 | 37 |
| 1 | 195 | 197 | -2 |
| 2 | 173 | 201 | -28 |
| 3 | 134 | 135 | -1 |
| 4 | 41 | 47 | -6 |
| Total | 4,738 | 4,738 |  |

## Estimates of Covid-19 infection

We now derive estimates of the rate of infection from this model of symptoms. Ideally the incidence of positive tests for each of the sums of symptoms would be based on a larger sample than is available from the Covid-19 survey. But we can make some tentative estimates based on a logit regression model relating the outcome of a positive test to the number of symptoms. Table 6 shows predicted infection rates by number of symptoms (*q*(*n*)) and their standard errors, and also the mean probabilities in the sample for different values for the number of symptoms (mean of *P*(*n*|*X*)) and their standard errors. The mean *P*(*n*|*X*) form weights for the infection rates *q*(*n*) to calculate the average infection rate in the population (∑ *P*(*n*|*X*)*q*(*n*)), which we estimate to be 0.047. Using the delta method to estimate the standard errors of *q*(*n*)*P*(*n*|*X*))^6^, the standard error of the mean infection rate is 0.0116, and the 95% confidence interval is between 0.024 and 0.070. Until we have more data on testing, it is not possible to obtain more precise estimates.

**Table 6:**
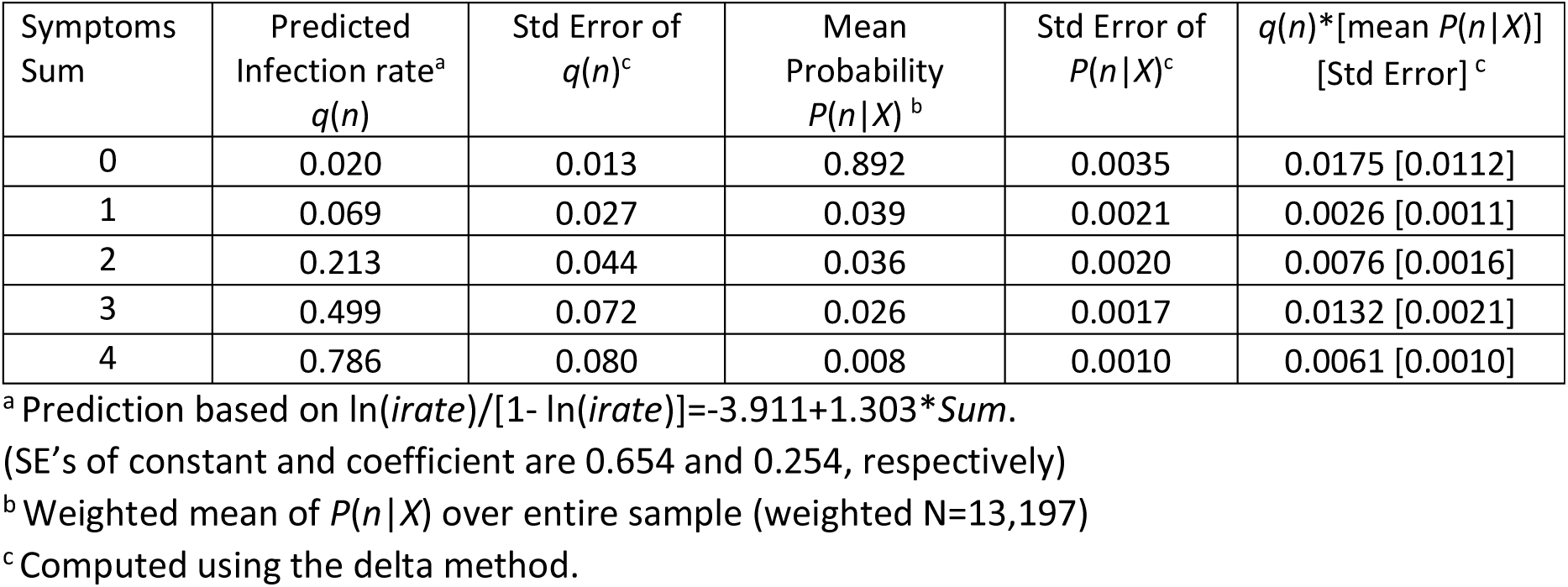
Calculation of Infection Rates Weighted by Probabilities of Number of Symptoms

These are estimates for the private household population only. The omission of nursing homes is likely to be important. The mean estimate of 0.047 and its confidence interval are close to the estimates by Hill Kulu and Peter Dorey (*Infection Rates from Covid-19 in Great Britain by Geographical Units: A Model-based Estimation from Mortality Data*, ESRC Centre for Population Change, University of St. Andrews 2020). They estimate that between 5 and 6% of people in Great Britain were infected by Covid-19 by the last third of April 2020, and state that ‘It is unlikely that the infection rate was lower than 3% or higher than 12%.’

Our estimate of a 4.7% infection rate (95% CI: 2.4% to 7%) during the period up to the end of April 2020 is consistent with the recent *Coronavirus (COVID-19) Infection Survey pilot: England, 17 July 2020*: *Initial data from the COVID-19 Infection Survey* (delivered in partnership with IQVIA, Oxford University and UK Biocentre), which indicates that between 26 April and 8 July, 6.3% of people in the community in England tested positive for antibodies against SARS-CoV-2 on a blood test, suggesting they had the infection in the past.

An advantage of the approach taken here is that we can calculate the infection rate for different values of *X*. Table 7 provides an example for education level, showing the mean *P(n*|*X)* for each of the four education levels, and also *q*(*n*) from Table 6. The last row indicates the estimated mean infection rate for each education level (∑ *P*(*n*|*X*)*q*(*n*)), and its standard error. We see it increases steadily with education level.

**Table 7:**
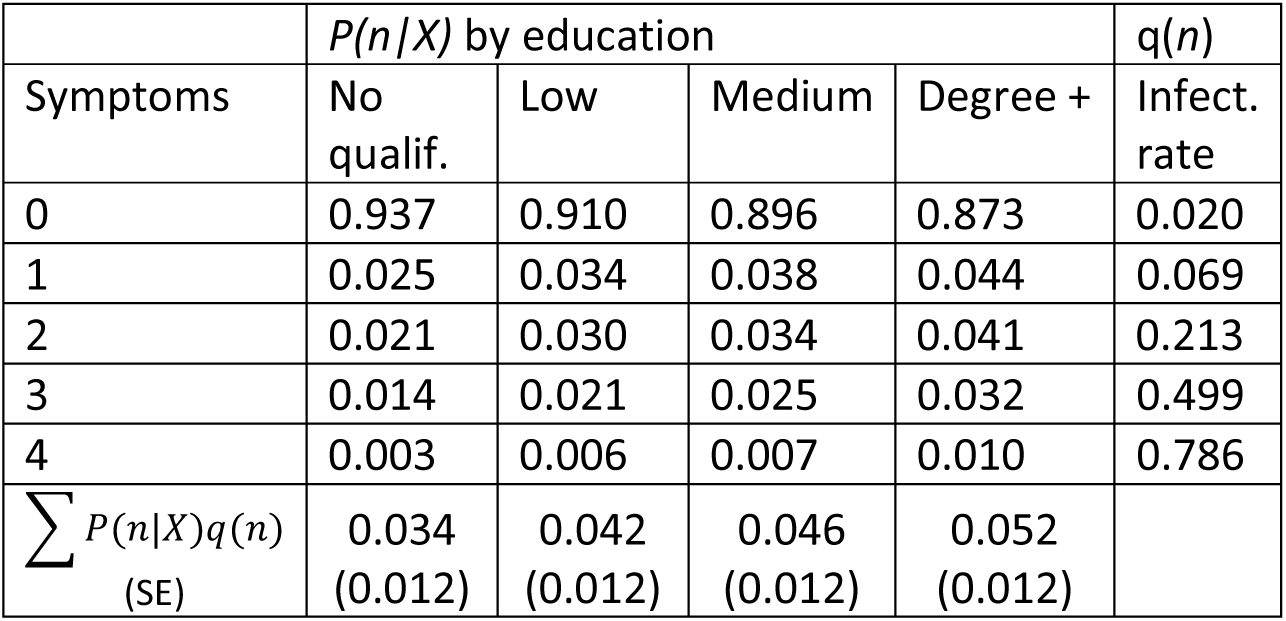
Symptom probabilities and estimated infection rate by Education level

Estimates at the regional level are consistent with findings of Kulu and Dorey (*ibid*.), namely London stands out with the highest infection rate (0.057) and infection rates increase with population density. Figure 1 shows regional infection rate estimates using an analogous computation procedure to that used in Table 7 (London is omitted because of its very higher density: 5,666 per square metre). The correlation between regional population density and the infection rate is 0.28. For comparison, the correlation between population density and the number of confirmed Covid-19 cases per 100,000 population (i.e. had a positive test) up to 29 April 2020 across 122 upper tier local authorities in England is also 0.28.^7^ Of course, our survey estimates assume that *q*(*n*), which translates symptoms into infection, does not vary with population density or other regional characteristics.

**Figure 1:**
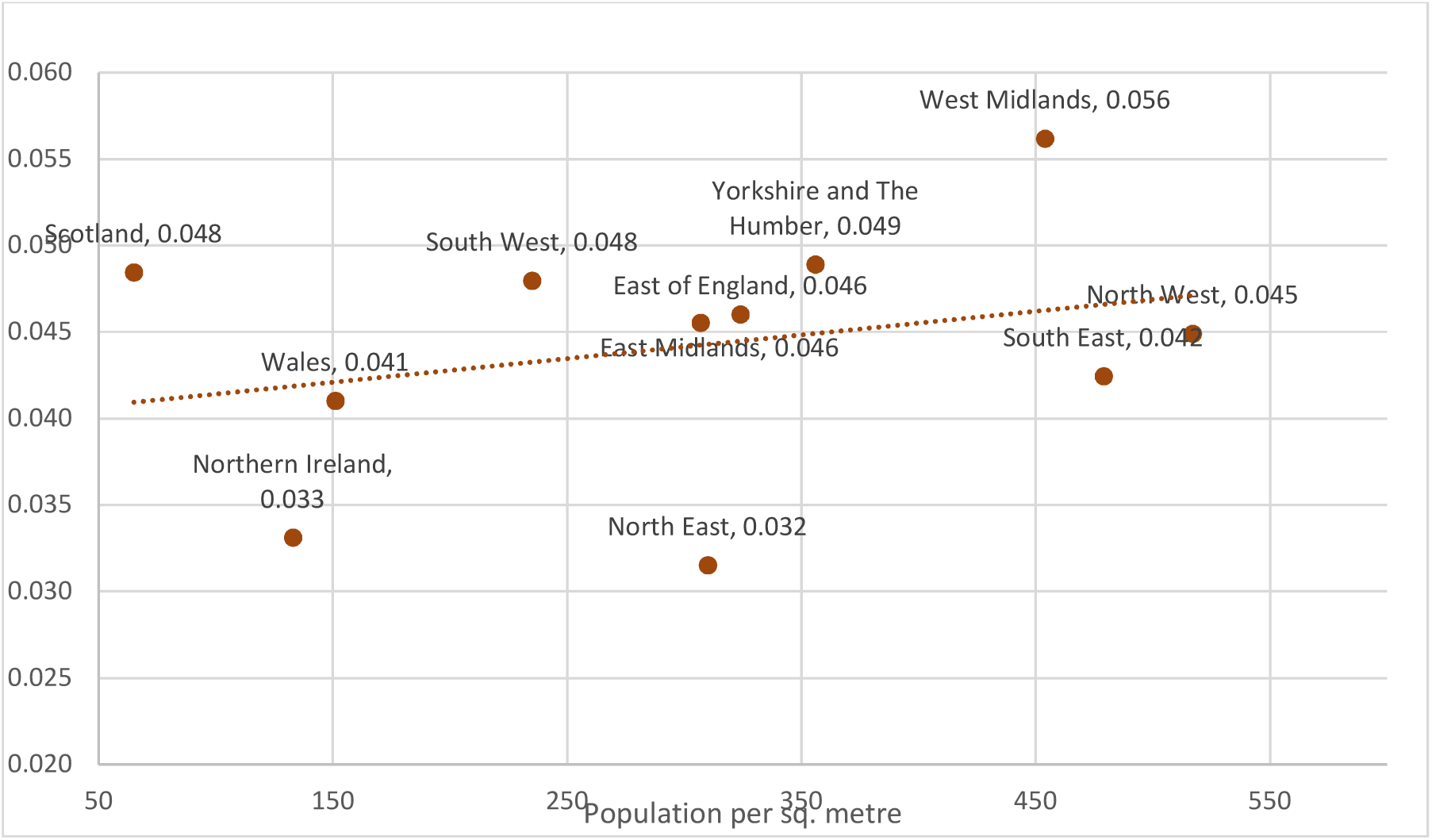
Mean predicted infection rate and population density, rho=0.28

Using the same estimation procedure for ∑ *P*(*n*|*X*)*q*(*n*) by age group, Figure 2 illustrates the decline in the mean infection rate with age. Recall from Table 1 that the estimates at the younger ages are based on parameter estimates for the relationship between age group and the latent index and these were not estimated with much precision for those aged less than 55.

**Figure 2:**
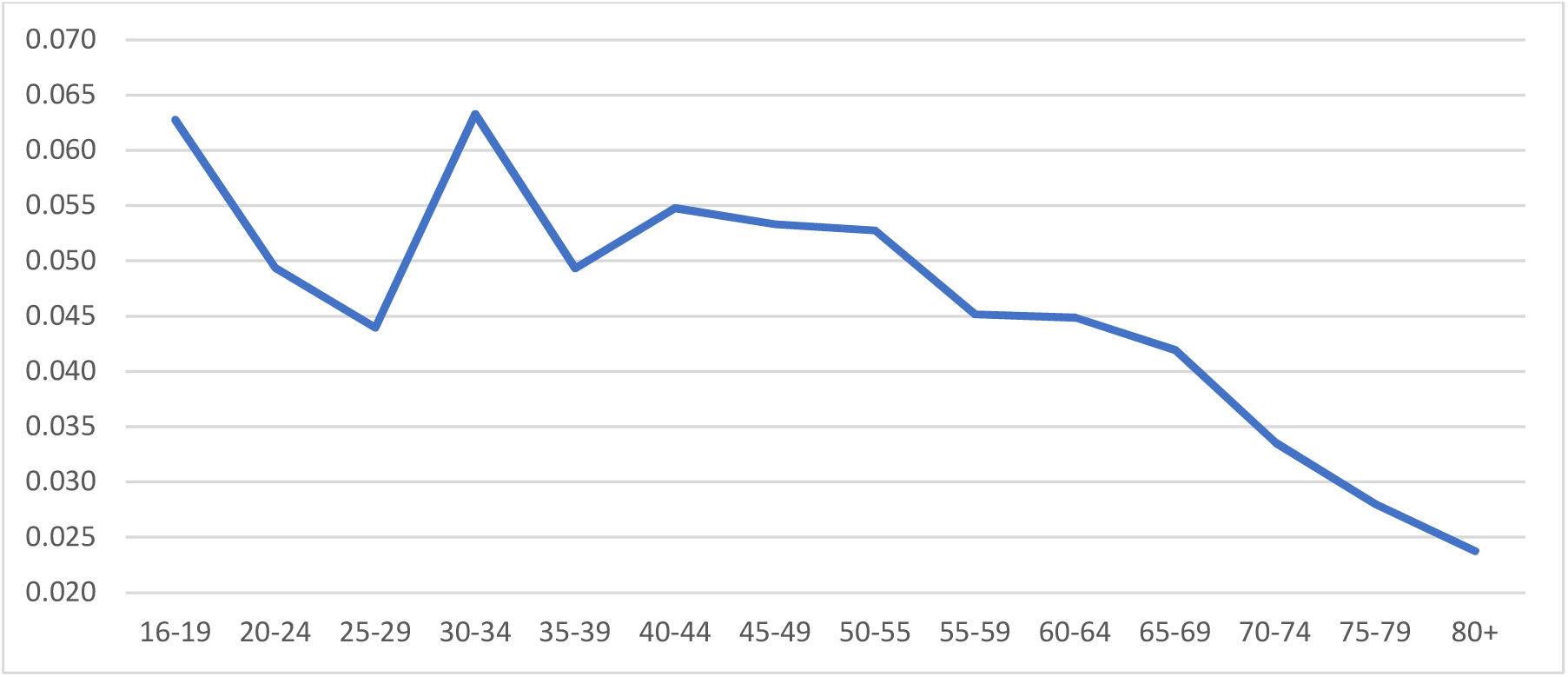
Mean predicted infection rate by Age group

## Alternative estimates of infection from ONS COVID-19 Infection Survey

The estimates above converting symptoms to the probability of infection are based on a small sample whose test results are known. We can also combine the *Understanding Society* survey data with estimates from a survey carried out by ONS (*Coronavirus (COVID-19) infections in the community in England: July 2020. Characteristics of people testing positive for the coronavirus (COVID-19) in England from the COVID-19 Infection Survey* (7 July)). It finds that among those reporting specifically having a cough, or fever, or loss of taste or smell on the day of testing, 10.1% (95% CI: 5.9% to 15.9%) tested positive for COVID-19 during 26 April to 27 June 2020, compared with an estimate of 0.28% ((95% CI: 0.22% to 0.34%) of those who did not report having these specific symptoms on the day of their positive test.^8^

In the *Understanding Society* data, 9.2% (SE=0.33%, 95% CI: 8.55% to 9.84%) reported at least one of these three symptoms. Combining this estimate of symptoms incidence with the infection rates during 26 April to 27 June 2020 from the ONS survey, the *Understanding Society* sample would suggest that 1.2% of the sample would test positive. But the sustained downward trend in new confirmed cases since 10 May strongly suggests that infection rates were much higher in the period leading up to 24-30 April, when the *Understanding Society* survey was carried out, than in May and June, when the ONS survey took place.

In the sample of those tested in the *Understanding Society* data, 34.5% of those reporting one of the three systems tested positive (compared with 10.1% in the ONS data), and there were 1.6% positive tests among those not reporting any of the three symptoms (compared with 0.3% in the ONS data). If we applied this relationship in the *Understanding Society* data, then we estimate a mean infection rate of 4.6%. This is very similar to that obtained earlier using the sum of four symptoms, which included fatigue as well as the three symptoms upon which ONS focuses, but the confidence interval is wider (1.7% to 7.6%) than in our earlier estimates, probably because our specification using number of symptoms contains more information.

## Alternative specifications of P(n|X) and q(n)

We consider two other specifications of *P*(*n*|*X*) and *q(n)* which differ in the number of symptoms considered in the two functions. The models for *q(n)* for in columns 2 and 5 of Table 8 indicate that the ‘new continuous cough’ symptom is not a very good predictor of a positive test in the survey data. Thus, in the model in column 4 we confine the range of *n* to the count of the three other symptoms *(sum3*). In contrast, the model in column 3 adds another symptom, ‘shortness of breath or trouble breathing’ to the count of symptoms (*sum5*). Table 9 shows the confidence interval using these two alternative models as well as our original one (*sum4*).

**Table 8:** Logit Models of Test Result and common Covid-symptoms (N=147) (Standard errors below coefficient estimate)

| Variable | 1 | 2 | 3 | 4 | 5 |
| --- | --- | --- | --- | --- | --- |
| *sum3*<sup>a</sup> |  |  |  | 2.009 | 2.055 |
|  |  |  |  | 0.387 | 0.391 |
| *sum4*<sup>b</sup> | 1.303 |  |  |  |  |
|  | 0.254 |  |  |  |  |
| *sum5*<sup>c</sup> |  |  | 1.003 |  |  |
|  |  |  | 0.193 |  |  |
| High temperature |  | 1.773 |  |  |  |
|  |  | 0.578 |  |  |  |
| New continuous cough |  | -0.489 |  |  | -0.444 |
|  |  | 0.550 |  |  | 0.541 |
| Fatigue |  | 2.546 |  |  |  |
|  |  | 0.834 |  |  |  |
| Loss of taste/smell |  | 1.973 |  |  |  |
|  |  | 0.565 |  |  |  |
| Constant | -3.91 | -4.54 | -3.67 | -4.49 | -4.35 |
|  | 0.65 | 0.89 | 0.59 | 0.78 | 0.79 |
| psuedo R-sq | 0.2981 | 0.4024 | 0.2837 | 0.3937 | 0.3983 |

<sup>a</sup>Number of the following symptoms: High temperature, Fatigue, Loss of taste or smell.
<sup>b</sup>Number of the following symptoms: High temperature, Persistent Cough, Fatigue, Loss of taste or smell.
<sup>c</sup>Number of the following symptoms: High temperature, Persistent Cough, Fatigue, Loss of taste or smell, Trouble breathing.

**Table 9:** Mean infection rate and confidence intervals

| Variable | Lower<br>95% limit | Mean<br>(SE) | Upper<br>95% limit |
| --- | --- | --- | --- |
| *sum3*<sup>a</sup> | 0.023 | 0.040<br>(0.008) | 0.056 |
| *sum4*<sup>b</sup> | 0.024 | 0.047<br>(0.012) | 0.070 |
| *sum5*<sup>c</sup> | 0.026 | 0.052<br>(0.013) | 0.077 |

<sup>a</sup>Number of the following symptoms: High temperature, Fatigue, Loss of taste or smell.
<sup>b</sup>Number of the following symptoms: High temperature, Persistent Cough, Fatigue, Loss of taste or smell.
<sup>c</sup>Number of the following symptoms: High temperature, Persistent Cough, Fatigue, Loss of taste or smell, Shortness of breath or trouble breathing.

The patterns of variation in *P*(*n*|*X*) with the different elements of *X*, such as age, education, etc., are similar to those in Table 4. The main difference in the mean infection rate comes from the different ‘*sums*’ in the *q(n)* functions. Using *sum3* produces a lower mean infection rate and a tighter confidence interval than the either *sum4* or *sum5*, but *sum3* was chosen as a possible specification through ‘pre-testing’, and no cross-validation was possible with the small sample available. This suggests that the confidence intervals are wider than the estimated standard errors would indicate. It is probably suitable cautious to say that the infection rate during the period up to the end of April 2020 was between 2% and 8%.

## Conclusions

The analysis of the new *Understanding Society COVID-19* survey indicates that the chances of Covid-19 infection increase with a person’s education level, are lower and declining with age among those aged over 55, and were higher in the West Midlands and London and lower in the North East than the rest of the country, and tended to increase with regional population density. There is also evidence that the infection rate was lower among those of a Caribbean origin. Our primary estimates suggest that the mean cumulative infection rate in the UK population living in private households was 0.047 during the period up to the end of April 2020, with a 95% confidence interval of between 0.024 and 0.070. This estimate is consistent with the finding that 6.3% of people in the community in England tested positive for antibodies against Covid-19 on a blood test between 26 April and 8 July, suggesting they had the infection in the past.

## Data Availability

The data used in the article are available to non-commerical users from the UK Data Service

## Footnotes

1 Institute for Social and Economic Research (2020) Understanding Society COVID-19 User Guide. Version 1.0, May 2020. Colchester: University of Essex.

2 Analysis of the reports of test results in relation to symptoms from the Covid-19 survey suggests we might also consider dropping ‘a new continuous cough’ from the count of number of symptoms, but our main results are based on all four symptoms mentioned above.

3 Analysis at the end of the paper (Table 8) indicates that the symptoms high temperature, loss of taste or smell, and fatigue have large impacts on the odds of testing positive, while a new continuous cough does not increase those odds.

4 We also estimated count data models (Poisson and Negative Binomial). Although producing broadly similar patterns for the impacts of the elements of *X*, in the crossvalidation exercise described below these models performed poorly outside of the estimation sample.

5 Equivalent household income is long-term average income divided by the square root of household size.

6 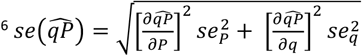

7 For the logs of these two variables, the correlation is 0.46 across the 122 upper tier local authorities and 0.40 across the standard England regions plus Wales.

8 Estimates are based on the first positive swab test for anyone testing positive in the study and the last negative swab test for anyone who never had a positive test. There was also a longer list of symptoms which respondents were asked to report: fever, muscle ache (myalgia), fatigue (weakness or tiredness), sore throat, cough, shortness of breath, headache, nausea/vomiting, abdominal pain, diarrhoea, loss of taste or loss of smell. Among those reporting any of these symptoms, 4.32% (95% CI: 2.87% to 6.22%) tested positive on a swab test. By comparison, the infection rate for those not reporting any symptoms was 0.25% (95% CI: 0.20% to 0.31%). l.

